# Motor Imagery-based Neurofeedback Using Visual, Auditory, Vibrotactile, and Proprioceptive Senses: A Randomized Controlled Trial

**DOI:** 10.1101/2025.08.27.25334613

**Authors:** Shun Sawai, Shin Murata, Shoya Fujikawa, Ryosuke Yamamoto, Keisuke Shima, Hideki Nakano

## Abstract

Motor imagery (MI)-based neurofeedback (NFB) has recently been shown to improve MI ability using visual, auditory, vibrotactile, and proprioceptive modalities; however, no study has compared all four of these sensory modalities to each other and to a control group. This randomized controlled trial aimed to examine the effects of MI-based NFB on MI ability (the ability to improve one’s motor abilities through visualizing movements) using visual, auditory, vibrotactile, and proprioceptive modalities. Fifty healthy young participants were recruited for this study and were randomly assigned to the control group or an NFB group using either visual, auditory, vibrotactile, or proprioceptive sensory feedback. All participants completed a pre-training evaluation of 20 trials without NFB. Next, 60 trials of the MI task (imagining themselves performing maximal dorsiflexion of the wrist joint) were performed during the training phase. After training, all groups were again assessed without NFB for 20 trials. For each NFB group, EEG was measured during the MI task, and event-related desynchronization (ERD) in motor-related areas was fed back to the participants in real time. The pre- and post-training results of ERD and subjective MI vividness were compared for each group; the results showed that ERD significantly increased after training in all four groups that performed NFB (p < 0.05) and that MI vividness increased significantly after training in all five groups (p < 0.05). The present study demonstrated that MI-based NFB using visual, auditory, vibrotactile, and proprioceptive modalities improved MI ability.

## 1 Introduction

Motor imagery (MI) is a cognitive process that helps a person to imagine being in motion without actual movement (Lotze et al., 2006; Olsson et al., 2010), and it has been shown to improve performance and promote motor learning (MarcIntyre et al., 2018). Functional brain imaging studies have also shown that MI induces brain activity similar to the activity produced during actual exercise or while observing exercise, indicating that MI has similar neural processes to those of actual exercise (Hanakawa et al., 2008). In addition, the dorsolateral frontal cortex and frontoparietal networks are characterized by motor-imagery-specific brain activity (Hardwick et al., 2018); this activity is associated with cognitive loading of executive preparation (Donohue et al., 2008) and working memory (Webler et al., 2022), the processes needed to hold mentally imagined movements (Kim et al., 2018). Thus, because MI activates brain regions related to real movement and cognitive processes, it has been used to train athletes with high-performance demands (Ridderinkhof et al., 2015) and to rehabilitate patients with neurological diseases (García Carrasco et al., 2013; Fujikawa et al., 2022). However, MI does not provide feedback to participants on whether their performance is good or bad, and individuals can experience different degrees of effectiveness for MI (van der Meulen et al., 2012). Recently, neurofeedback (NFB) technology, which provides feedback on brain activity during MI, has been developed as a tool to solve this problem (Riahi et al., 2022; Power et al., 2020).

NFB can promote MI and improve motor performance (Riahi et al., 2022; Hwang et al., 2009). The NFB is a closed-loop technique that provides real-time feedback on brain activity to participants to help them control voluntary brain activity (Girges et al., 2022). In particular, MI-based NFB uses event-related desynchronization (ERD) in the μ-wave band of the primary motor cortex (M1) (Pineda et al., 2003; Enomae et al., 2017). Many previous studies have shown that the M1 is activated by MI (Hanakawa et al., 2008; Hardwick et al., 2018), causing μ ERD (Takemi et al., 2013; Francuz et al., 2011); therefore, this activity indicates the degree of MI. Furthermore, MI-based NFB has been applied to the rehabilitation of patients with neurological diseases such as stroke (Mihara et al., 2021) or Parkinson’s disease (Cuomo et al., 2022) and has been shown to improve physical function.

MI-based NFB can be administered through visual (Zapala et al., 2018; Tinaz et al., 2022), auditory (Nakano et al., 2018), tactile (Grigorev et al., 2021), and proprioceptive modalities (Darvishi et al., 2017; Irimia et al., 2017), all of which have been reported to improve MI ability. Therefore, MI-based NFB using several sensory modalities has been shown to be effective. In addition, previous studies comparing sensory modalities have shown that MI-based NFB using proprioceptive stimuli is more effective than using visual stimuli. Furthermore, a comparison of MI-based NFB using visual and auditory stimuli reported that visual stimulation is more effective for acquiring motor skills, whereas auditory stimulation is more effective for retaining motor skills. Therefore, comparing sensory modalities in MI-based NFB has contributed to the development of more effective MI-based NFB. However, previous studies have only compared the effectiveness of MI-based NFB using one or two sensory modalities; a comparison involving all four sensory modalities has not yet been conducted. Therefore, this study aimed to compare the effectiveness of MI-based NFB using visual, auditory, vibrotactile, and proprioceptive modalities in a randomized controlled trial. Identifying the optimal sensory modality for MI-based NFB may contribute to developing more effective MI-based NFB.

## 2 Methods

### 2.1 Participants

Fifty healthy young men who had never received MI-based NFB (age: 20.42 ± 0.67 years, height: 172.78 ± 5.79 cm, body weight: 65.86 ± 8.23 kg) were recruited for the study. Patients with a history of motor or cognitive dysfunction or those regularly taking medication, including psychotropic drugs, were excluded. The Edinburgh Handedness Inventory (Oldfield et al., 1971) was used to confirm that all participants were right-handed. This study was conducted in accordance with the Declaration of Helsinki, and informed consent was obtained from all participants. This study was approved by the Research Ethics Committee of Kyoto Tachibana University (approval no. 22-49).

### 2.2 Sample Size Calculation

Using the G*Power software (Faul et al., 2007), the sample size was determined as 40 to obtain an effect size of 0.30, α = 0.05, and power (1-β) = 0.80 at a confidence level of 95%.

### 2.3 Study Protocol

This was a randomized controlled trial. Randomization was performed using computer-generated sequences. In the pre-training evaluation phase, participants performed 20 consecutive trials of the MI task without NFB. All MI tasks in this study consisted of 5 s of rest followed by 5 s of MI, repeated as one trial. Participants were then categorized into five groups: a control group (n = 10, age: 20.40 ± 0.70 years, height: 173.20 ± 8.03 cm, body weight: 68.00 ± 10.43 kg), visual FB group (n = 10, age: 20.40 ± 0.70 years, height: 173.70 ± 4.50 cm, body weight: 64.40 ± 5.42 kg), auditory FB group (n = 10, age: 20.50 ± 0.71 years, height: 172.90 ± 7.01 cm, body weight: 67.20 ± 12.42 kg), vibrotactile FB group (n = 10, age: 20.20 ± 0.42 years, height: 173.80 ± 4.39 cm, body weight: 66.40 ± 5.19 kg), and proprioceptive FB group (n = 10, age: 20.60 ± 0.84 years, height: 170.30 ± 4.47 cm, body weight: 63.30 ± 5.70 kg). In the training phase, the MI task was repeated for 60 consecutive trials. The control group performed the MI task without NFB, whereas participants in the NFB groups used the assigned sensory modalities to receive real-time feedback regarding their brain activity during the task. Finally, participants performed 20 consecutive trials of the MI task without NFB during the post-training evaluation phase. The pre- and post-training ERDs and subjectively reported MI vividness during the MI task were compared (Figure 1).

**Figure 1.**
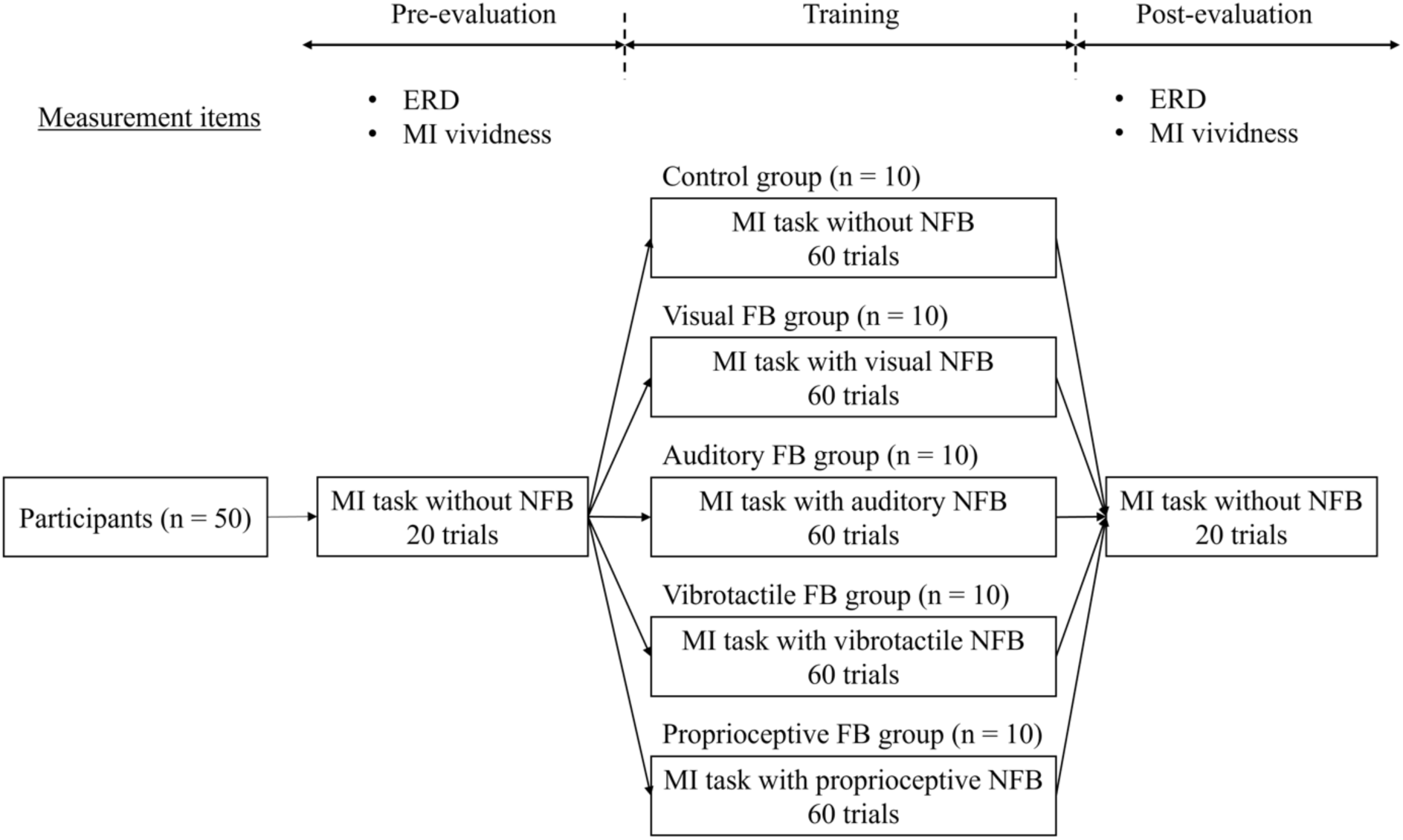
Study protocol. First, participants performed 20 trials of the MI task without NFB, and ERD and MI vividness were evaluated during the MI task. Next, participants were randomly divided into five groups: control group, visual FB group, auditory FB group, vibrotactile FB group, and proprioceptive FB group. The control group performed the MI task without NFB, while the other groups performed the MI task with NFB using the assigned sensory modality during the training phase. Finally, 20 trials of the MI task without NFB were performed, and ERD and MI vividness during the MI task were evaluated again. ERD: event-related desynchronization; MI: motor imagery; FB: feedback; NFB: neurofeedback.

### 2.4 Motor Imagery

The participants sat in a chair with a backrest, placed both hands on a desk, and imagined themselves performing a dorsiflexion movement of the left wrist joint at 100% of the maximum voluntary contraction intensity from a kinesthetic perspective (Figure 2). Because kinesthetic motor imagery has been reported to produce μ ERD more effectively than visual motor imagery (Neuper et al., 2005), all participants in this study were instructed to perform kinesthetic motor imagery.

**Figure 2.**
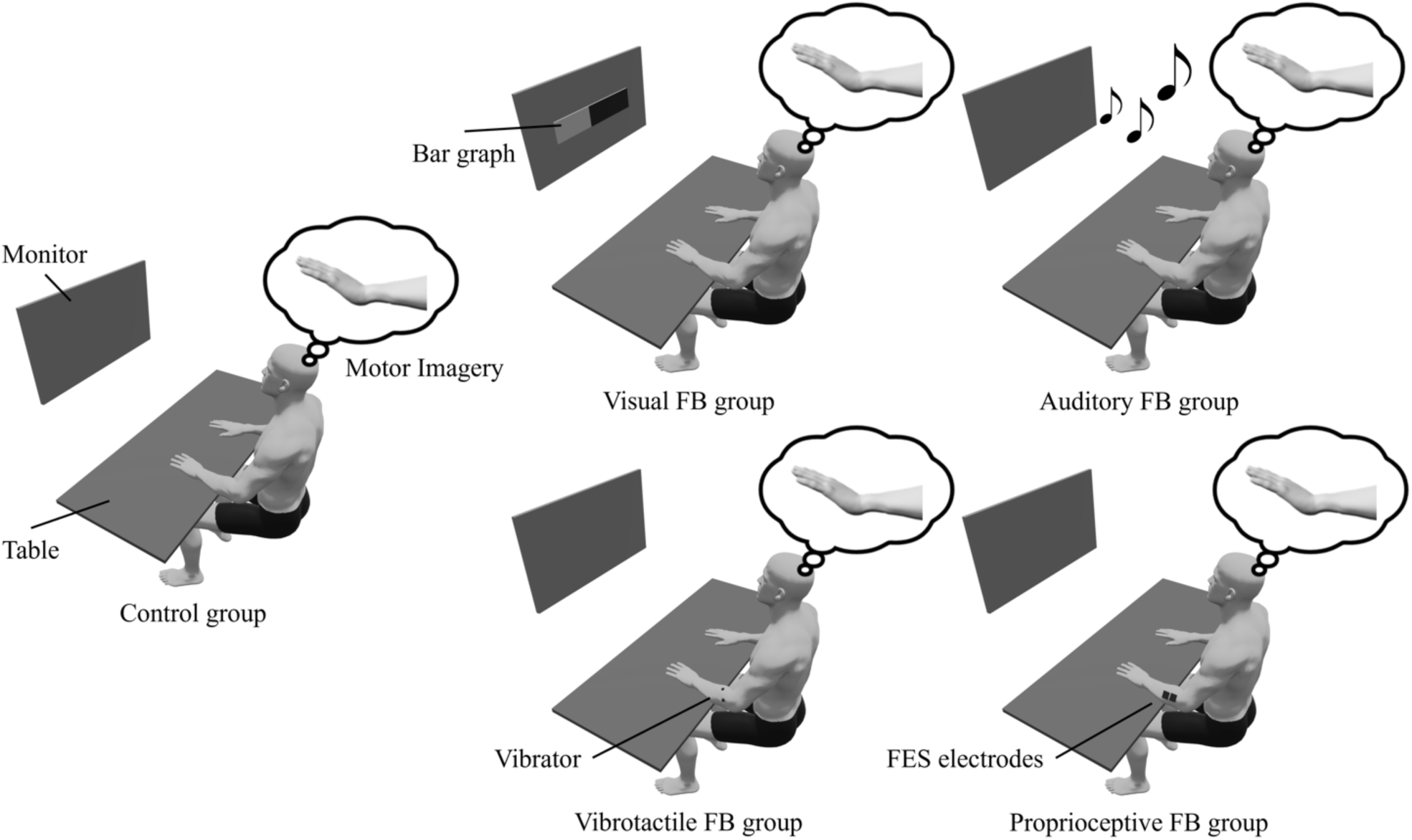
Experimental setup. The participant sat in a chair with a backrest and placed both hands on a desk. Participants were instructed to watch a monitor during the MI task. In the visual FB group, ERD was expressed as an increase or decrease in a bar graph. In the auditory FB group and the vibrotactile FB group, ERD was represented by sound and vibration frequencies. In the proprioceptive FB group, ERD was expressed by joint movement using electrical stimulation. ERD: event-related desynchronization; MI: motor imagery; FB: feedback; FES: functional electrical stimulation.

### 2.5 Neurofeedback

During the training phase, EEG was measured during the MI task, and the μ ERD of M1 was fed back in real time to the participants assigned to each NFB group but not to the control group. EEG was measured using an electroencephalograph (EEG-9100; Nihon Kohden Corp., Tokyo, Japan) and an active dry electrode system (Miyuki Giken Co., Ltd., Tokyo, Japan). EEG signals were recorded by 19 channels (Fp1, Fp2, F7, F3, Fz, F4, F8, T3, C3, Cz, C4, T4, T5, P3, Pz, P4, T6, O1, and O2) according to the international 10-20 system, and reference electrodes were placed on both auricular surfaces. The sampling rate was set to 1,000 Hz. The recorded EEG data were analyzed using Microsoft Visual Studio (Microsoft Corp., Redmond, WA, USA). Frequency analysis was used to calculate ERD in the μ-wave band (8-13 Hz) of the sensorimotor area (C4). The time window for calculating ERD was set at 100 ms. The ERD *E*(*t*) at time *t* was calculated by Equation (1), using the resting wave activity *R*_*rest*_ and wave activity during the imagery task *R*_*ima*(*e*_(*t*) (Pfurtscheller et al., 1999).

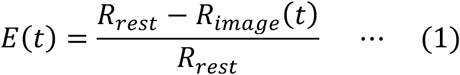

Here, ERD indicates a decrease in μ-wave activity during MI when compared to the resting state. Therefore, *E*(*t*) can take values from -∞ to 1, with ERD falling between 0 and 1. ERD in the μ-wave band of C4 is also associated with motor imagery (Pineda et al., 2003; Enomae et al., 2017).

Different sensory modalities were used to deliver the ERD feedback in each group. In the visual FB group, a bar graph was displayed on a monitor, and ERD was expressed as an increase or decrease in the graph. In the auditory FB group, ERD was represented by a change in musical pitch: the higher the ERD, the higher the frequency and pitch of the sound stimulus. The sound level was set at 50 dB and generated by a speaker placed in front of the participants. Before training, participants were presented with the highest pitch, B4 (493 Hz), and the lowest pitch, C4 (261 Hz), to confirm they could hear both. The frequency of the sound stimulation *F*(*t*) at time *t* is shown in Equation (2), using the ERD value *E*(*t*), the threshold of ERD *E*_*th*_, the maximum frequency *F*_*max*_, and minimum frequency *F*_*min*_ of the sound stimulation. In this study, *F*_*max*_ was 493 and *F*_*min*_ was 261.

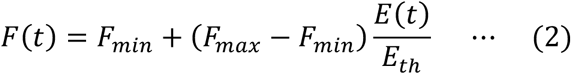

In the vibrotactile FB group, a vibrating device (LD14-002; Nidec Components Corp., Tokyo, Japan) was attached over the left extensor carpi radialis longus, and the ERD was expressed by the frequency of the vibration stimulation; the higher the ERD, the higher the frequency of the vibration stimulation applied. The vibrating device measured 14 mm × 11.2 mm × 2.6 mm and was a linear resonant actuator with an output frequency range of 120–180 Hz. Before the training phase, vibration stimuli at 120 Hz and 180 Hz were applied to confirm they were perceived. The frequency of the vibration *V*(*t*) at time *t* is given by Equation (3), using the ERD value *E*(*t*), the threshold of ERD *E*_*th*_, the minimum frequency of the vibration stimulation *V*_*min*_, and the vibration coefficient *K*.

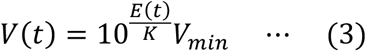

Here, *V*_*min*_denotes the minimum perceived frequency. *K* is shown in equation (4) using the maximum of ERD *E*_*max*_, the maximum frequency *V*_*max*_, and the minimum frequency *V*_*min*_ of vibration stimulation. In this study, *V*_*max*_ was 180 Hz and *V*_*min*_ was 120 Hz.

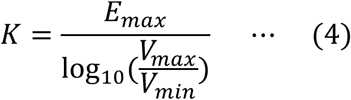

In the proprioceptive FB group, electrical stimulation was applied to the extensor carpi radialis longus using an electrical stimulator (SEN-3401; Nihon Kohden Corp., Tokyo, Japan), and ERD was expressed as the resulting dorsiflexion movement of the left wrist joint; the higher the ERD, the greater the dorsiflexion movement of the wrist joint induced by electrical stimulation. The current of electrical stimulation *I*(*t*) at time *t* is given by Equation (5), using the ERD value *E*(*t*), the ERD threshold *E*_*th*_, and the maximum current amount *I*_*max*_ of the electrical stimulation.

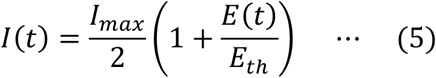

Here, *I*_*max*_ is the amount of current that induces dorsiflexion of the wrist joint into the final position. No adverse events, such as skin injury due to electrical stimulation, occurred in the proprioceptive FB group.

### 2.6 Evaluation

ERD and MI vividness were measured during the MI task. The ERD value was measured by Equation 1, and the average of the maximum values for each trial were used in the analysis. MI vividness was evaluated using a paper-based visual analog scale (VAS) (Moriuchi et al., 2020; Altemira et al., 2023); participants self-evaluated the vividness of the motor images by marking a 100-mm horizontal line, where “0 = not at all” and “100 = very vivid image.”

### 2.7 Statistical Analysis

The Shapiro-Wilk test was first used to confirm the normality of all data. A one-way analysis of variance was then used to compare age, height, and weight between the groups. ERD and VAS were compared using a two-factor repeated-measures analysis of variance, with the two factors of group (control, visual FB, auditory FB, vibrotactile FB, and proprioceptive FB) and time (pre, post). The Bonferroni method was applied to the post-hoc tests. SPSS ver. 28.0 (IBM Corp., Armonk, NY, USA) was used for all statistical analyses, and statistical significance was defined at p < 0.05.

## 3 Results

There were no significant differences in age (F = 0.47, p = 0.76, partial η^2^ = 0.04), height (F = 0.59, p = 0.67, partial η^2^ = 0.05), or body weight (F = 0.55, p = 0.70, partial η^2^ = 0.05) between participant groups. ERD and VAS results are shown in Tables 1 and 2. Comparison of ERD showed no time × group significant interaction (F = 1.07, p = 0.38, partial η^2^ = 0.09), but revealed a significant main effect for time (F = 36.56, p < 0.05, partial η^2^ =0.45). Post-test results showed a significant increase in ERD after training for all NFB groups (p < 0.05, Figure 3). A comparison of VAS scores also showed no significant time × group interaction (F = 1.03, p = 0.40, partial η^2^ = 0.08), but revealed a significant main effect for time (F = 83.00, p < 0.05, partial η^2^ = 0.65). The post-training evaluation results of all five groups showed a significant increase in the VAS scores after training (p < 0.05, Figure 4).

**Figure 3.**
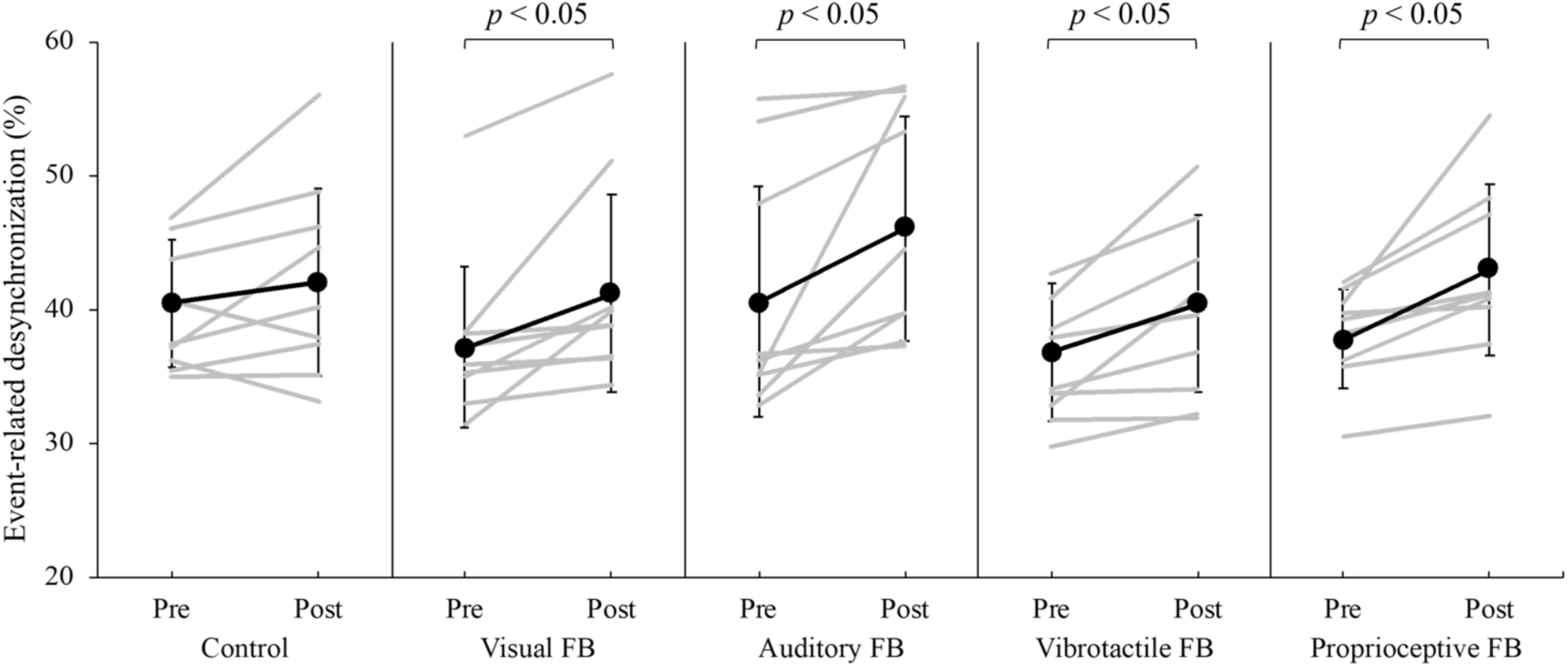
Changes in ERD of each group before and after training. The gray lines show the data for each participant, and the black line with endpoints shows the mean value for each group. Error bars indicate the standard deviation. The visual FB group, auditory FB group, vibrotactile FB group, and proprioceptive FB group showed a significant increase in ERD after training (p < 0.05), whereas the control group showed no change in ERD before and after training (p > 0.05). ERD: event-related desynchronization; FB: feedback.

**Figure 4.**
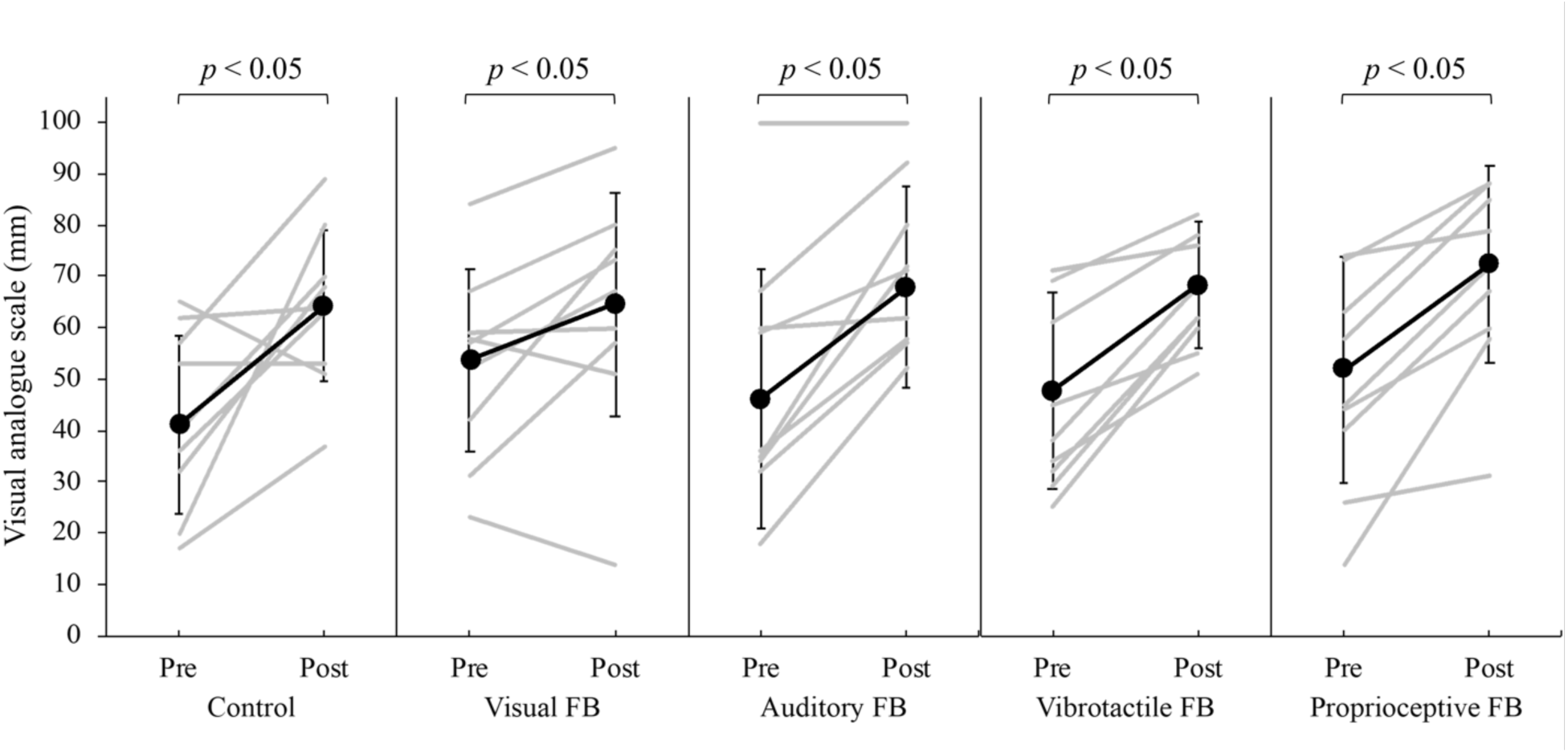
Changes in VAS rating of each group before and after training. The gray lines show data for each participant, and the black line with endpoints shows the mean value for each group. Error bars indicate the standard deviation. All groups reported significantly higher VAS ratings of image vividity after training (p < 0.05). VAS: visual analog scale; FB: feedback.

**Table 1.** μ ERD values pre- and post-training.

|  | Pre (%) |  | Post (%) |  |
| --- | --- | --- | --- | --- |
|  | Mean | SD | Mean | SD |
| Control | 40.45 | 4.81 | 41.97 | 6.98 |
| Visual FB | 37.18 | 5.97 | 41.18 | 7.33 |
| Auditory FB | 40.55 | 8.62 | 46.05 | 8.43 |
| Vibrotactile FB | 40.96 | 6.48 | 45.47 | 8.30 |
| Proprioceptive FB | 37.73 | 3.69 | 43.00 | 6.41 |

ERD: event-related desynchronization; SD: standard deviation; FB: feedback.

**Table 2.** VAS values pre- and post-training.

|  | Pre (mm) |  | Post (mm) |  |
| --- | --- | --- | --- | --- |
|  | Mean | SD | Mean | SD |
| Control | 41.10 | 17.27 | 64.10 | 14.75 |
| Visual FB | 53.60 | 17.75 | 64.50 | 21.71 |
| Auditory FB | 46.20 | 25.15 | 67.80 | 19.54 |
| Vibrotactile FB | 47.90 | 19.20 | 68.30 | 12.45 |
| Proprioceptive FB | 51.80 | 21.86 | 72.40 | 19.29 |

VAS: visual analogue scale; SD: standard deviation; FB: feedback.

## 4 Discussion

This randomized controlled trial aimed to examine the effects of MI-based NFB using visual, auditory, vibrotactile, and proprioceptive modalities on MI ability. The results showed that ERD of M1, which indicates the degree of MI, significantly increased after training in all groups (visual, auditory, vibrotactile, and proprioceptive) that received NFB. The subjective evaluation of MI vividness also increased significantly in all groups after training. The results show that MI-based NFB improves MI ability compared with no NFB.

### 4.1 The Effect of MI-based NFB on μ ERD

In motor learning, feedback on knowledge of results (KR) enables error learning and further improves performance (Newell et al., 1976; Salmoni et al., 1984). In MI, feedback from brain activity provides the KR and can improve MI ability by promoting the voluntary control of MI-related brain activity (Pineda et al., 2003; Enomae et al., 2017). In this study, no change was observed in ERD before and after training in the control group; however, ERD increased after training in all groups in which brain activity was fed back into a sensory modality. The MI ability of the NFB groups may have improved because the ERD feedback enabled error learning. Previous studies have shown the effectiveness of MI-based NFB (Takemi et al., 2013; Francuz et al., 2011; Hwang et al., 2009; Marcos-Martínez et al., 2021; Dettmers et al., 2016; Grigorev et al., 2021), and the increase of ERD in the NFB groups seen in this study is consistent with these previous studies.

The study found no significant main effects of group or significant group-by-time interactions in the μ ERD comparisons. This result indicates that the effect of NFB does not vary significantly depending on the sensory modality used in MI-based NFB. Previously, the effects of MI-based NFB using visual (Zapala et al., 2018; Tinaz et al., 2022), auditory (Nakano et al., 2018), vibrotactile (Grigorev et al., 2021), and proprioceptive (Darvishi et al., 2017; Irimia et al., 2017) FBs have been investigated separately. Additionally, studies comparing these sensory modalities have shown that NFB is more effective with vibrotactile FB than visual FB (Shabani et al., 2021; Vukelić et al., 2015). It has also been reported that auditory FB is more effective in retaining acquired motor skills than visual FB, whereas visual FB is more effective in acquiring motor skills than auditory FB (Ronsse et al., 2011). These previous studies consistently reported improved MI ability with NFB compared to no NFB. In the present study, the control group showed no significant μ ERD improvement before and after training, whereas the four groups receiving NFB showed significant μ ERD improvement after training. These findings suggest that, in MI-based NFB, the presence or absence of feedback may influence MI ability improvement more than the sensory modality providing the feedback. However, based on previous studies that found differences in NFB effects by sensory modality, the effects of MI-based NFB may be maximized by selecting the most appropriate sensory modality based on the phase of training or skill acquisition and the type of motor task. A possible reason why the effects of MI-based NFB did not differ across sensory modalities in this study is the dominance of attention focus on the sensory modality. Previous studies reported that the optimal attention focus that improves performance is different for each individual (Sakurada et al., 2016; Sakurada et al., 2022; Sawai et al., 2022). Therefore, the dominance of attention focus for each sensory modality during MI-based NFB may have obscured differences in MI ability by modality.

### 4.2 The Effect of MI-based NFB on MI Vividness

In contrast, MI vividness significantly increased after training in all groups. MI vividness has been reported to increase with repetitions of the MI task (Iso et al., 2021), but no effect on MI vividness was observed for the combination of NFB, electrical stimulation, and behavioral observations for MI repetition (Ono et al., 2013; Sawai et al., 2022). In this study, MI vividness increased after training in all groups, regardless of the presence or absence of NFB or the sensory modality of the feedback. Therefore, this study’s results suggest that MI vividness is not influenced by the presence or absence of NFB but rather by the repetition of MI to improve MI ability.

### 4.3 Limitation

This study had some limitations. First, because this study was conducted as a randomized controlled trial, not all sensory FB conditions could be applied to every participant. Sensory modalities that enhance the effects of NFB may provide an attention-focus advantage for each individual. Therefore, future studies should use a crossover design to test the effects of MI-based NFB across all sensory modalities within the same participants. Second, we did not examine the effects of MI-based NFB on motor imagery tasks with varying difficulty levels. Previous studies have reported increased activity in motor-related areas during MI tasks of higher difficulty (Taube et al., 2015). Therefore, it is necessary to examine whether MI ability can be improved using NFB with MI tasks of different difficulty levels. Third, this study did not examine the long-term retention effects of MI-based NFB on improvement in MI ability. It has been reported that the retention effect of learning is particularly high when using auditory FB (Ronsse et al., 2011). Future studies should examine the retention effects of learning for each sensory modality to clarify the long-term characteristics of sensory modalities in MI-based NFB. Fourth, the participants in this study were healthy young men. Unifying participant demographics reduces confounding factors such as age and gender, but may limit the generalizability of the study’s results. Future research should incorporate a variety of participants. Fifth, EMG was not measured during the MI task in this study. Therefore, it cannot be ruled out that muscle activity occurred during the MI task and that only MI was performed. In future studies, it is recommended to measure EMG to isolate the effect of NFB on MI alone, completely excluding muscle activity. Sixth, this study could not be completely double-blinded. Participants received MI-based NFB through a randomly assigned sensory modality and were unaware of the specific stimuli used beyond the modality they received. However, the researchers knew which sensory modality was assigned to each participant to deliver the corresponding neurofeedback stimulus. It is important to implement more precise blinding procedures in future studies. Seventh, this study performed MI-based NFB with four different sensory stimuli but did not include a random NFB group. Therefore, the independent effects of providing NFB cannot be elucidated in this study. Eighth, this study could not verify whether the four NFB sensory modalities were of equal ease of use or saliency. Therefore, it is possible that the cognitive load during NFB differed among the four sensory modalities. Future studies are needed to validate these results by controlling for equivalent cognitive load and to identify NFB sensory modalities with lower cognitive demands. Ninth, this study used the ERD of C4 as an objective measure of MI accuracy to validate the effectiveness of MI-based NFB. However, advanced analyses, such as whole-brain region analyses or functional connectivity, were not performed. In future studies, additional EEG analyses should be conducted to verify the effects of MI-based NFB from multiple perspectives.

## 6 Author Contributions

HN: conceptualization. HN: methodology. SS and HN: formal analysis. SS, SF, RY, and HN: investigation. KS, and HN: resources. SS and HN: data curation. SS and HN: writing and preparation of the original draft. SS, SM, SF, RY, KS, and HN: draft review and editing. SS and HN: data visualization. SM and HN: supervision. SM and HN: project administration. HN: funding acquisition. All authors contributed to the manuscript revision and have read and approved the submitted version.

## 8 Abbreviation list

EEG: electroencephalography
ERD: event-related desynchronization
KR: knowledge of results
MI: motor imagery
NFB: neurofeedback
VAS: visual analog scale.

## 9 Acknowledgments

We would like to thank all the volunteers who participated in this study. We thank Editage [http://www.editage.com] for English language editing and reviewing of the manuscript.

## 11 Data Availability Statement

The data supporting the findings of this study are available from the corresponding author upon reasonable request. The data are not publicly available because they contain information that could compromise the privacy of research participants.

## References

Altemira, G.H., Skiba, N., Lécuyer, A., Bougrain, L., Fleck, S. (2023). “Multisensory neurofeedback design for KMI embodiment.”, in Proceedings of the 2023 ACM International Conference on Interactive Media Experiences Workshops (IMXw ‘23). (New York: Association for Computing Machinery), 56–58. doi: 10.1145/3604321.3604352.

Cuomo, G., Maglianella, V., Ghanbari Ghooshchy, S., Zoccolotti, P., Martelli, M., Paolucci, S. et al. (2022). Motor imagery and gait control in Parkinson’s disease: techniques and new perspectives in neurorehabilitation. Expert Rev Neurother. 22, 43–51. doi: 10.1080/14737175.2022.2018301. [Epub 2021 Dec 28]. PMID: 34906019.

Darvishi, S., Gharabaghi, A., Boulay, C.B., Ridding, M.C., Abbott, D., Baumert, M. (2017). Proprioceptive Feedback Facilitates Motor Imagery-Related Operant Learning of Sensorimotor β- Band Modulation. Front Neurosci. 11, 60. doi: 10.3389/fnins.2017.00060, PMID: 28232788, PMCID: PMC5299002.

Dettmers, C., Braun, N., Büsching, I., Hassa, T., Debener, S., Liepert, J. (2016). Neurofeedback-based motor imagery training for rehabilitation after stroke. Nervenarzt. 87, 1074–1081. doi: 10.1007/s00115-016-0185-y, PMID: 27573884.

Donohue, S.E., Wendelken, C., Bunge, S.A. (2008). Neural correlates of preparation for action selection as a function of specific task demands. J Cogn Neurosci. 20, 694–706. doi: 10.1162/jocn.2008.20042, PMID: 18052782.

Enomae, T., Fujimori, H., Tanaka, H. (2017). Analysis of the Switching Information on Motor Imagery for Asynchronous BCI. International Journal of Affective Engineering. 16, 131–137. doi: 10.5057/ijae.IJAE-D-16-00019.

Faul, F., Erdfelder, E., Lang, A.G., Buchner, A. (2007). G*Power 3: a flexible statistical power analysis program for the social, behavioral, and biomedical sciences. Behav Res Methods. 39, 175–191. doi: 10.3758/bf03193146, PMID: 17695343.

Francuz, P., Zapała, D. (2011). The suppression of the μ rhythm during the creation of imagery representation of movement. Neurosci Lett. 495, 39–43. doi: 10.1016/j.neulet.2011.03.031. [Epub 2011 Mar 22]. PMID: 21406213.

Fujikawa, S., Ohsumi, C., Ushio, R., Tamura, K., Sawai, S., Yamamoto, R., et al. (2022). “Potential Applications of Motor Imagery for Improving Standing Posture Balance in Rehabilitation”, in Neurorehabilitation and Physical Therapy, ed. Nakano, H. (London: IntechOpen), 27–42.

García Carrasco, D., Aboitiz Cantalapiedra, J. (2016). Effectiveness of motor imagery or mental practice in functional recovery after stroke: a systematic review. Neurologia. 31, 43–52. doi: 10.1016/j.nrl.2013.02.003. [Epub 2013 Apr 17]. PMID: 23601759.

Gäumann, S., Gerber, R.S., Suica, Z., Wandel, J., Schuster-Amft, C. (2021). A different point of view: the evaluation of motor imagery perspectives in patients with sensorimotor impairments in a longitudinal study. BMC Neurol. 21, 297. doi: 10.1186/s12883-021-02266-w, PMID: 34315411, PMCID: PMC8314460.

Girges, C., Vijiaratnam, N., Zrinzo, L., Ekanayake, J., Foltynie, T. (2022). Volitional Control of Brain Motor Activity and Its Therapeutic Potential. Neuromodulation. 25, 1187–1196. doi: 10.1016/j.neurom.2022.01.007. [Epub 2022 Feb 28]. PMID: 35241365.

Grigorev, N.A., Savosenkov, A.O., Lukoyanov, M.V., Udoratina, A., Shusharina, N.N., Kaplan, A.Y. et al. (2021). A BCI-Based Vibrotactile Neurofeedback Training Improves Motor Cortical Excitability During Motor Imagery. IEEE Trans Neural Syst Rehabil Eng. 29, 1583–1592. doi: 10.1109/TNSRE.2021.3102304. [Epub 2021 Aug 13]. PMID: 34343094.

Hanakawa, T., Dimyan, M.A., Hallett, M. (2008). Motor planning, imagery, and execution in the distributed motor network: a time-course study with functional MRI. Cereb Cortex. 18, 2775–2788. doi: 10.1093/cercor/bhn036. [Epub 2008 Mar 20]. PMID: 18359777, PMCID: PMC2583155.

Hardwick, R.M., Caspers, S., Eickhoff, S.B., Swinnen, S.P. (2018). Neural correlates of action: Comparing meta-analyses of imagery, observation, and execution. Neurosci Biobehav Rev. 94, 31–44. doi: 10.1016/j.neubiorev.2018.08.003. [Epub 2018 Aug 9]. PMID: 30098990.

Hwang, H.J., Kwon, K., Im, C.H. (2009). Neurofeedback-based motor imagery training for brain-computer interface (BCI). J Neurosci Methods. 179, 150–156. doi: 10.1016/j.jneumeth.2009.01.015. [Epub 2009 Jan 29]. PMID: 19428521.

Irimia, D.C., Cho, W., Ortner, R., Allison, B.Z., Ignat, B.E., Edlinger, G. et al. (2017). Brain-Computer Interfaces With Multi-Sensory Feedback for Stroke Rehabilitation: A Case Study. Artif Organs. 41, E178–E184. doi: 10.1111/aor.13054, PMID: 29148137.

Iso, N., Moriuchi, T., Fujiwara, K., Matsuo, M., Mitsunaga, W., Hasegawa, T. et al. (2021). Hemodynamic Signal Changes During Motor Imagery Task Performance Are Associated With the Degree of Motor Task Learning. Front Hum Neurosci. 15, 603069. doi: 10.3389/fnhum.2021.603069, PMID: 33935666, PMCID: PMC8081959.

Kim, Y.K., Park, E., Lee, A., Im, C.H., Kim, Y.H. (2018). Changes in network connectivity during motor imagery and execution. PLoS One. 13, e0190715. doi: 10.1371/journal.pone.0190715, PMID: 29324886, PMCID: PMC5764263.

Lotze, M., Halsband, U. (2006). Motor imagery. J Physiol Paris. 99, 386–395. doi: 10.1016/j.jphysparis.2006.03.012. [Epub 2006 May 22]. PMID: 16716573.

MacIntyre, T.E., Madan, C.R., Moran, A.P., Collet, C., Guillot, A. (2018). Motor imagery, performance and motor rehabilitation. Prog Brain Res. 240, 141–159. doi: 10.1016/bs.pbr.2018.09.010. [Epub 2018 Oct 24]. PMID: 30390828.

Marcos-Martínez, D., Martínez-Cagigal, V., Santamaría-Vázquez, E., Pérez-Velasco, S., Hornero, R. (2021). Neurofeedback Training Based on Motor Imagery Strategies Increases EEG Complexity in Elderly Population. Entropy (Basel). 23, 1574. doi: 10.3390/e23121574, PMID: 34945880, PMCID: PMC8700498.

Mihara, M., Fujimoto, H., Hattori, N., Otomune, H., Kajiyama, Y., Konaka, K. et al. (2021). Effect of Neurofeedback Facilitation on Poststroke Gait and Balance Recovery: A Randomized Controlled Trial. Neurology. 96, e2587–e2598. doi: 10.1212/WNL.0000000000011989. [Epub 2021 Apr 20]. PMID: 33879597, PMCID: PMC8205450.

Moriuchi, T., Nakashima, A., Nakamura, J., Anan, K., Nishi, K., Matsuo, T. et al. (2020). The Vividness of Motor Imagery Is Correlated With Corticospinal Excitability During Combined Motor Imagery and Action Observation. Front Hum Neurosci. 14, 581652. doi: 10.3389/fnhum.2020.581652, PMID: 33088268, PMCID: PMC7500410.

Nakano, H., Kodama, T., Ukai, K., Kawahara, S., Horikawa, S., Murata, S. et al. (2018). Reliability and Validity of the Japanese Version of the Kinesthetic and Visual Imagery Questionnaire (KVIQ). Brain Sci. 8, 79. doi: 10.3390/brainsci8050079, PMID: 29724042, PMCID: PMC5977070.

Nakano, H., Kodama, T., Murata, S., Nakamoto, T., Fujihara, T., Ito, Y. (2018). Effect of Auditory Neurofeedback Training on Upper Extremity Function and Motor Imagery Ability in a Stroke Patient: A Single Case Study. Int J Clin Res Trials. 3, 126. doi: 10.15344/2456-8007/2018/126.

Nakano, H., Tachibana, M., Fujita, N., Sawai, S., Fujikawa, S., Yamamoto, R. et al. (2022). Reliability and validity of the Japanese movement imagery questionnaire-revised second version. BMC Res Notes. 15, 334. doi: 10.1186/s13104-022-06220-y, PMID: 36284354, PMCID: PMC9594881.

Neuper, C., Scherer, R., Reiner, M., Pfurtscheller, G. (2005). Imagery of motor actions: differential effects of kinesthetic and visual-motor mode of imagery in single-trial EEG. Brain Res Cogn Brain Res. 25, 668–677. doi: 10.1016/j.cogbrainres.2005.08.014. [Epub 2005 Oct 19]. PMID: 16236487.

Newell, K.M. (1976). Knowledge of results and motor learning. Exerc Sport Sci Rev. 4, 195–228. PMID: 798689.

Oldfield, R.C. (1971). The assessment and analysis of handedness: the Edinburgh inventory. Neuropsychologia. 9, 97–113. doi: 10.1016/0028-3932(71)90067-4, PMID: 5146491.

Olsson, C.J., Nyberg, L. (2010). Motor imagery: if you can’t do it, you won’t think it. Scand J Med Sci Sports. 20, 711–715. doi: 10.1111/j.1600-0838.2010.01101.x, PMID: 20338003.

Ono, T., Kimura, A., Ushiba, J. (2013). Daily training with realistic visual feedback improves reproducibility of event-related desynchronisation following hand motor imagery. Clin Neurophysiol. 124, 1779–1786. doi: 10.1016/j.clinph.2013.03.006. [Epub 2013 May 3]. PMID: 23643578.

Pfurtscheller, G., Lopes da Silva, F.H. (1999). Event-related EEG/MEG synchronization and desynchronization: basic principles. Clin Neurophysiol. 110, 1842–1857. doi: 10.1016/s1388-2457(99)00141-8, PMID: 10576479.

Pineda, J.A., Silverman, D.S., Vankov, A., Hestenes, J. (2003). Learning to control brain rhythms: making a brain-computer interface possible. IEEE Trans Neural Syst Rehabil Eng. 11, 181–184. doi: 10.1109/TNSRE.2003.814445, PMID: 12899268.

Power, L., Neyedli, H.F., Boe, S.G., Bardouille, T. (2020). Efficacy of low-cost wireless neurofeedback to modulate brain activity during motor imagery. Biomed Phys Eng Express. 6, 035024. doi: 10.1088/2057-1976/ab872c, PMID: 33438669.

Riahi, N., Ruth, W., D’Arcy, R.C.N., Menon, C. (2022). A Method for Using Neurofeedback to Guide Mental Imagery for Improving Motor Skill. IEEE Trans Neural Syst Rehabil Eng. 31, 131–138. doi: 10.1109/TNSRE.2022.3218514, PMID: 36318564.

Ridderinkhof, K.R., Brass, M. (2015). How Kinesthetic Motor Imagery works: a predictive-processing theory of visualization in sports and motor expertise. J Physiol Paris. 109, 53–63. doi: 10.1016/j.jphysparis.2015.02.003. [Epub 2015 Mar 25]. PMID: 25817985.

Ronsse, R., Puttemans, V., Coxon, J.P., Goble, D.J., Wagemans, J., Wenderoth, N. et al. (2011). Motor learning with augmented feedback: modality-dependent behavioral and neural consequences. Cereb Cortex. 21, 1283–1294. doi: 10.1093/cercor/bhq209. [Epub 2010 Oct 28]. PMID: 21030486.

Sakurada, T., Hirai, M., Watanabe, E. (2016). Optimization of a motor learning attention-directing strategy based on an individual’s motor imagery ability. Exp Brain Res. 234, 301–311. doi: 10.1007/s00221-015-4464-9. [Epub 2015 Oct 14]. PMID: 26466828.

Sakurada, T., Yoshida, M., Nagai, K. (2022). Individual Optimal Attentional Strategy in Motor Learning Tasks Characterized by Steady-State Somatosensory and Visual Evoked Potentials. Front Hum Neurosci. 15, 784292. doi: 10.3389/fnhum.2021.784292, PMID: 35058765, PMCID: PMC8763707.

Salmoni, A.W., Schmidt, R.A., Walter, C.B. (1984). Knowledge of results and motor learning: a review and critical reappraisal. Psychol Bull. 95, 355–386. PMID: 6399752.

Sawai, S., Fujikawa, S., Murata, S., Abiko, T., Nakano H. (2022). Dominance of Attention Focus and Its Electroencephalogram Activity in Standing Postural Control in Healthy Young Adults. Brain Sci. 12, 538. doi: 10.3390/brainsci12050538, PMID: 35624924, PMCID: PMC9138695.

Sawai, S., Fujikawa, S., Ushio, R., Tamura, K., Ohsumi, C., Yamamoto, R. (2022). Repetitive Peripheral Magnetic Stimulation Combined with Motor Imagery Changes Resting-State EEG Activity: A Randomized Controlled Trial. Brain Sci. 12, 1548. doi: 10.3390/brainsci12111548, PMID: 36421872, PMCID: PMC9688706.

Shabani, F., Nisar, S., Philamore, H., Matsuno, F. (2021). Haptic vs. Visual Neurofeedback for Brain Training: A Proof-of-Concept Study. IEEE Trans Haptics. 14, 297–302. doi: 10.1109/TOH.2021.3077492. [Epub 2021 Jun 17]. PMID: 33945486.

Takemi, M., Masakado, Y., Liu, M., Ushiba, J. (2013). Event-related desynchronization reflects downregulation of intracortical inhibition in human primary motor cortex. J Neurophysiol. 110, 1158–1166. doi: 10.1152/jn.01092.2012. [Epub 2013 Jun 12]. PMID: 23761697.

Taube, W., Mouthon, M., Leukel, C., Hoogewoud, H.M., Annoni, J.M., Keller, M. (2015). Brain activity during observation and motor imagery of different balance tasks: an fMRI study. Cortex. 64, 102–114. doi: 10.1016/j.cortex.2014.09.022. [Epub 2014 Oct 27]. PMID: 25461711.

Tinaz, S., Kamel, S., Aravala, S.S., Elfil, M., Bayoumi, A., Patel, A. et al. (2022). Neurofeedback-guided kinesthetic motor imagery training in Parkinson’s disease: Randomized trial. Neuroimage Clin. 34, 102980. doi: 10.1016/j.nicl.2022.102980. [Epub 2022 Mar 2]. PMID: 35247729, PMCID: PMC8897714.

van der Meulen, M., Allali, G., Rieger, S.W., Assal, F., Vuilleumier, P. (2014). The influence of individual motor imagery ability on cerebral recruitment during gait imagery. Hum Brain Mapp. 35, 455–470. doi: 10.1002/hbm.22192. [Epub 2012 Sep 27]. PMID: 23015531, PMCID: PMC6869533.

Vukelić, M., Gharabaghi, A. (2015). Oscillatory entrainment of the motor cortical network during motor imagery is modulated by the feedback modality. Neuroimage. 111, 1–11. doi: 10.1016/j.neuroimage.2015.01.058. [Epub 2015 Feb 7]. PMID: 25665968.

Webler, R.D., Fox, J., McTeague, L.M., Burton, P.C., Dowdle, L., Short, E.B. et al. (2022). DLPFC stimulation alters working memory related activations and performance: An interleaved TMS-fMRI study. Brain Stimul. 15, 823–832. doi: 10.1016/j.brs.2022.05.014. [Epub 2022 May 27]. PMID: 35644517.

Zapała, D., Francuz, P., Zapała, E., Kopiś, N., Wierzgała, P., Augustynowicz, P. et al. (2018). The Impact of Different Visual Feedbacks in User Training on Motor Imagery Control in BCI. Appl Psychophysiol Biofeedback. 43, 23–35. doi: 10.1007/s10484-017-9383-z, PMID: 29075937, PMCID: PMC5869881.

